# Prevalence and factors associated with post-natal growth restriction among low-birth-weight infants at a rural county referral hospital, Kenya: A retrospective observational study

**DOI:** 10.64898/2026.08.04.26359674

**Authors:** Lucy Lyanda, Roseline Ochieng, Alfred Keter

## Abstract

**Background:** The globally low birth weight is estimated to account for about 15-20% of all births. Global annual preterm delivery is about fifteen million, more than 60% of which occurs in Asia and Sub-Saharan Africa. The Kenyan preterm birth rate is estimated to be 14 per 100 live births. Prematurity, the leading cause of low birth weight, and its associated complications account for a large proportion of neonatal and under 5 morbidity and mortality. Premature and low birth weight (LBW) infants have rapid growth with increased nutritional demands despite their inadequate nutrient stores. This predisposes them to post-natal growth restriction (PNGR). Timely and adequate nutrition is important to prevent PNGR that has been shown to have long term neurodevelopmental and growth consequences. Immaturity and clinical instability of these infants makes providing adequate nutrition for them a great challenge. This study was done to determine the prevalence of and factors associated with PNGR among LBW infants at discharge from a rural public referral hospital in Kenya.

**Methods:** This was a single center retrospective descriptive study, of infants with birth weight <1800g discharged between 1^st^ January 2022 and 31^st^ December 2023, done to determine the prevalence of and the factors associated with PNGR among these infants at discharge. PNGR was defined as poor growth after birth during the initial hospitalization period. Cross sectional PNGR was discharge weight less than 10^th^ centile while longitudinal PNGR was a drop in centiles lines for weight at discharge. Moderate PNGR was discharge weight between 3^rd^ and 10^th^ centile while severe PNGR was discharge weight less than the 3^rd^ centile. Weight at birth and discharge was obtained and presented as Z scores and centiles using the respective gender specific intergrowth 21 charts. Maternal, clinical and nutritional data were also obtained from the patient files and analyzed to determine their association with PNGR.

**Results:** Among the 143 LBW infants included in the study, the prevalence of PNGR was 63% (n=90) (severe 44.8%) and 53.2% (n=76) (severe 16.1%) for cross sectional and longitudinal PNGR respectively. Statistically significant associations for longitudinal PNGR were found in patients who received any supplements aOR(95%CI) 3.0(1.1-8.3) p=0.037, took 14 days or more to regain birth weight aOR (95%CI) 0.1(0.1-0.4) p=<0.01 and were admitted for 30 days or more aOR(95%CI) 0.3(0.1-0.9) p=0.025.

**Conclusion:** More than half of LBW infants in Nyeri County Referral Hospital are discharged with PNGR, a sizeable proportion with severe form. There is need to update local LBW infants’ feeding protocols to include breast milk fortification, parenteral nutrition and avail resources to support the high nutritional needs of this population.

## Introduction

Prematurity is the leading cause of low birth weight (LBW) deliveries. Globally, the LBW rates are estimated to be around 15-20% of all live births. The World Health Organization (WHO) estimates the burden of premature deliveries to be at about 11% globally. Southeast Asia and Sub-Saharan Africa account for more than 60% of all preterm births (1). The Kenyan preterm birth rate is estimated to be 14 per 100 live births and the incidence of low birth weight is about 10% of all births. Prematurity and its associated complications remain a leading cause of under 5 morbidity and mortality. Preterm birth interferes with normal intrauterine third trimester nutrient accretion. Preterm and LBW infants, despite their inadequate nutrient stores, have rapid growth with consequent increased nutritional demands (3) and often fail to gain weight as expected from their intrauterine growth projections (4). Very preterm infants (born between 28- and 32-weeks’ gestation) have immature digestive systems and are often clinically unstable which renders timely and adequate nutrition difficult to achieve (2, 3). Special attention to nutrition of this infant population has been shown to improve and optimize growth during the initial neonatal critical care, the general hospital admission and beyond (5, 6). Poor in-hospital growth has long term neurodevelopmental and growth consequences including prolonged hospital stay and poor brain growth. The extremely low birth weight infants (birth weight less than 1000g) have been shown to have slower growth throughout childhood (5, 6). International guidelines including ESPHAGAN (7) and WHO all recommend that the preterm/LBW infant be fed with mothers’ own milk which should be fortified to meet the high nutritional demands of the preterm/LBW infant. The guidelines also recommend that before full enteral feeds are achieved, parenteral nutrition should be used to bridge the protein and calorie deficit. These guidelines are difficult to implement fully in low resource settings. Unfortunately, there are very few local guidelines on feeding premature and LBW infants and as such, much variability exists between units and practitioners (7, 8). There are many factors influencing the preterm and low birth weight infants’ growth during the initial admission after birth that have been studied (6, 12). Post natal growth restriction (PNGR) has been defined as either cross-sectional (weight <10^th^ centile at a specified time, mostly at term or at discharge) or longitudinal (drop in birth weight centile over a specified time mostly term or discharge). It is important to understand and optimize preterm and LBW infants’ growth during initial hospital stay due to both the short- and long-term health implications of post-natal growth restriction.

## Materials and methods

This was a retrospective descriptive study conducted at the Nyeri County Referral Hospital (NCRH) Newborn Unit (NBU), Kenya. This is a rural level 5 health facility, whose NBU has an official bed capacity of 38 and receives an average of 60 admissions monthly, approximately a third of which are due to low birth weight (LBW). All neonates with a birth weight <1800g, regardless of gestation at birth, who were admitted for at least seven days and discharged from the unit between 1^st^ January 2022, and 31^st^ December 2023 were the studied. We excluded admission beyond 72 hours after birth, death, discharge or transfer out before seven days of life. We also planned to exclude any neonate with a major congenital anomaly that may interfere with feeding and consequently growth like gastroschisis, major congenital heart diseases or major craniofacial anomalies but none were identified. During the study period, the feeding protocols were unchanged. This includes intravenous 10% dextrose on the day of birth, introduction of standard maintenance doses of sodium and potassium on the second day for all neonates and introduction of expressed breast milk (EBM) once available. Some neonates are given formula, if available, in case there is no EBM. Micronutrient supplements, iron, folate, and multivitamins are prescribed once infants achieve full enteral feeds at 180ml/kg/day. There is no practice of total parenteral nutrition or human milk fortification. Infants admitted to the unit are weighed every day using digital weighing scales. The weighing scales are always kept on a firm surface, and the infants are brought to the weighing area. The machines are recalibrated to zero before each weight measurement and they also undergo monthly routine maintenance and calibration by the biomedical department. Preterm and LBW infants who can feed orally and are clinically stable are discharged once they attain a weight of about 1800g for follow up in the outpatient clinic.

Fisher’s formula was used to calculate the sample size.

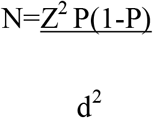

where N= sample size,

Z= level of confidence which for this study will be set at 95% (1.96),

P= expected prevalence- 91.2% (Based on a South African study (17),

d= precision set at 95%.

Based on the estimate of 15 LBW infant admissions to the unit per month, we estimated enrollment of 360 infants into the study which is well above the minimum sample size of 124.

220 patient records were identified from the Ministry of Health (MOH) admissions’ register, five files were excluded due to admission 72 hours after birth, 17 were discharged, while 40 had died before seven days. Fifteen files could not be traced, leaving 143 files that we used to collect our data.

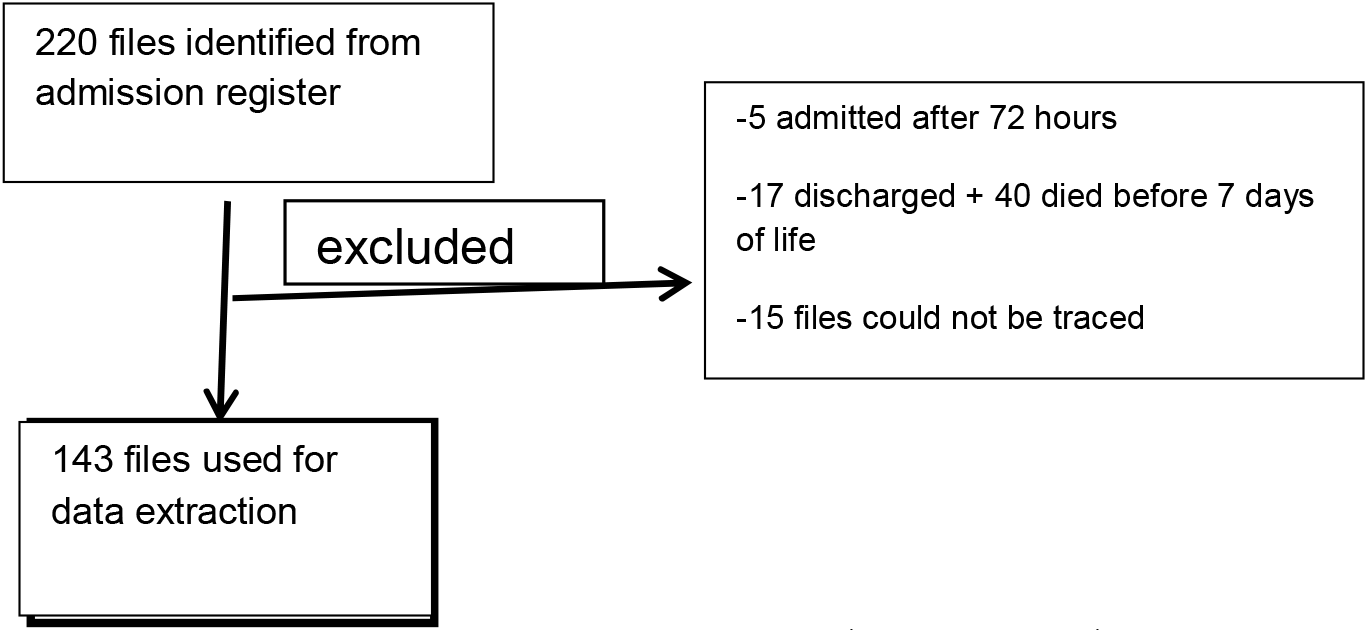

The patients’ files were accessible for research purpose from 10^th^ June to 20^th^ September 2024. A structured data collection tool was used to collect demographic, nutritional and clinical data from the identified patients’ files into REDCap (Research Electronic Data Capture) and later downloaded onto excel spreadsheets. Data was fully anonymized before analysis.

Demographic data of interest included time and date of birth, delivery method (cesarean or vaginal), date of admission, birth weight, gestation at birth, gender, date when birth weight was regained, date of discharge and weight at discharge.

Maternal data included mother’s age, parity, morbidities (gestational diabetes, gestational hypertension, pre-eclampsia, HIV and whether on antiretroviral therapy) and administration of antenatal corticosteroids.

Clinical data included Apgar score at 5 minutes, date of first file diagnosis during hospital stay of sepsis, file diagnosis of NEC, RDS, BPD, PDA, and anemia. We also extracted information regarding the use and duration of CPAP, any treatment with supplemental oxygen and any blood transfusion.

Nutritional data included date and time of enteral feed initiation, type of feed initiated, whether breast milk or formula, date of first attaining full enteral feeds (180ml/kg/day or no IVF prescribed), use of supplements (multivitamin, iron, folate), and date each supplement was started. The birth and discharge weights were plotted on the intergrowth 21^st^ appropriate charts and both the corresponding centiles and Z scores were recorded.

We derived the time in days, to achieve full enteral feeds (either 180ml/kg/day or no intravenous fluids prescribed), to regain birth weight and the duration of admission from the primary data extracted from the patient files.

Data was summarized into categorical variables expressed as frequencies and proportions, and continuous variables expressed as means with standard deviations (SD), or medians with interquartile ranges (IQR).

PNGR was defined two ways longitudinal PNGR (a drop from birth weight to discharge weight Z score by >1SD) and as cross-sectional PNGR (discharge weight <10^th^ centile).

PNGR was further classified as severe PNGR if discharge weight is at or below the 3^rd^ centile or a drop of 2SD or more from birth weight or moderate PNGR if discharge weight is between 3^rd^ and 10^th^ centile or a drop of 1-2SD from birth weight.

The prevalence of PNGR was calculated for both longitudinal and cross sectional as the proportion of infants meeting the respective definitions out of the total. Associations between PNGR and the independent variables were tested using Chi square test with significant association set at 5%. Binary logistic regression analysis was used to assess the association between PNGR and independent variables. The odd ratio with their 95% confidence interval was calculated. Statistical analysis was done using R version 4.4.3 (R core team 2025).

### Ethical considerations

The study was approved by the Aga Khan Institutional scientific and ethics review committee (ISERC) ISREC 2024/ISERC-24(v1) who waived informed consent for data access from medical records. A research permit Ref No. 891164 was also obtained from the National Commission for Science, Technology & Innovation (NACOSTI). Additional approvals were sought and granted from the Nyeri County department of health services and the Nyeri county referral hospital administration. Data collected was fully anonymized before the analysis and the data was stored in a password protected laptop only accessible to the principal investigator.

## Results

### Demographic characteristics of the LBW infants studied

Data was retrieved from 143 inpatient files of infants <1800g at birth who had been discharged from the newborn unit (NBU) at the Nyeri County Referral Hospital (NCRH) between January 2022 and December 2023 after an admission period of 7 days or more. Most of the infants were male (53.9%), had a median gestation of 32 weeks, (interquartile range (IQR): 29-34) and a range of 26-41 weeks. The mean birth weight was 1410g (standard deviation (SD): 266g) with a range of 635-1795g with 28.7% of the study subjects below the 10^th^ centile for birth weight. Most of the infants (85.9%) had an Apgar score ≥5 and the majority (79.7%) were born within the study site.

A summary of the infant characteristics is presented in Table 1 below.

**Table 1:** Demographic characteristics of the LBW infants n=143.

| Variable | Frequencies (%) | Mean (SD) /Median (IQR),<br>Range (Min-Max) |
| --- | --- | --- |
| **Sex** |  |  |
| Male | 77 (53.9) |  |
| Female | 66 (46.2) |  |
| **Gestation age at birth (weeks)** |  |  |
| < 28 | 14 (9.8) |  |
| 28-31 | 55 (38.5) |  |
| 32-35 | 57 (39.9) |  |
| ≥36 | 17 (11.9) |  |
| Median (IQR) |  |  |
| Range (Min-max) |  | 32 (29-34)<br>26 – 41 |
| **Birth weight (grams)** |  |  |
| <1000 | 12 (8.4) |  |
| 1000-1499 | 66 (46.2) |  |
| >1499 | 65 (45.5) |  |
| Mean (SD) |  | 1410 (266) |
| Range (Min-max) |  | 635 - 1795 |
| **Birth weight centiles** |  |  |
| <3<sup>rd</sup> | 29 (20.3) |  |
| 3<sup>rd</sup>-10<sup>th</sup> | 12 (8.4) |  |
| ≥10<sup>th</sup> | 102 (71.3) |  |
| **Birth weight Z scores (SD)** |  |  |
| <-2 | 26 (18.2) |  |
| -2 to -1 | 21 (14.7) |  |
| >-1 | 96 (67.1) |  |
| **Mode of delivery** |  |  |
| SVD | 113 (79.1) |  |
| CS | 30 (21) |  |
| **Multiple gestation \*** |  |  |
| Yes | 38 (26.6) |  |
| No | 105 (73.4) |  |
| **Apgar score at 5 minutes** |  |  |
| <5 | 15 (10.5) |  |
| ≥5 | 128 (89.5) |  |
| **Source of admission** |  |  |
| This facility | 114 (79.7) |  |
| Other facility | 24 (16.8) |  |
| Home | 5 (3.5) |  |

\*Not every baby in a multiple delivery was included in the study

### Demographic characteristics of the mothers of the LBW infants studied

The mothers had an age range of 13-43 years with a median of 26 years. The majority had no pregnancy comorbidity like gestational diabetes, PET and HIV. These characteristics are summarized in Table 2 below.

**Table 2:**
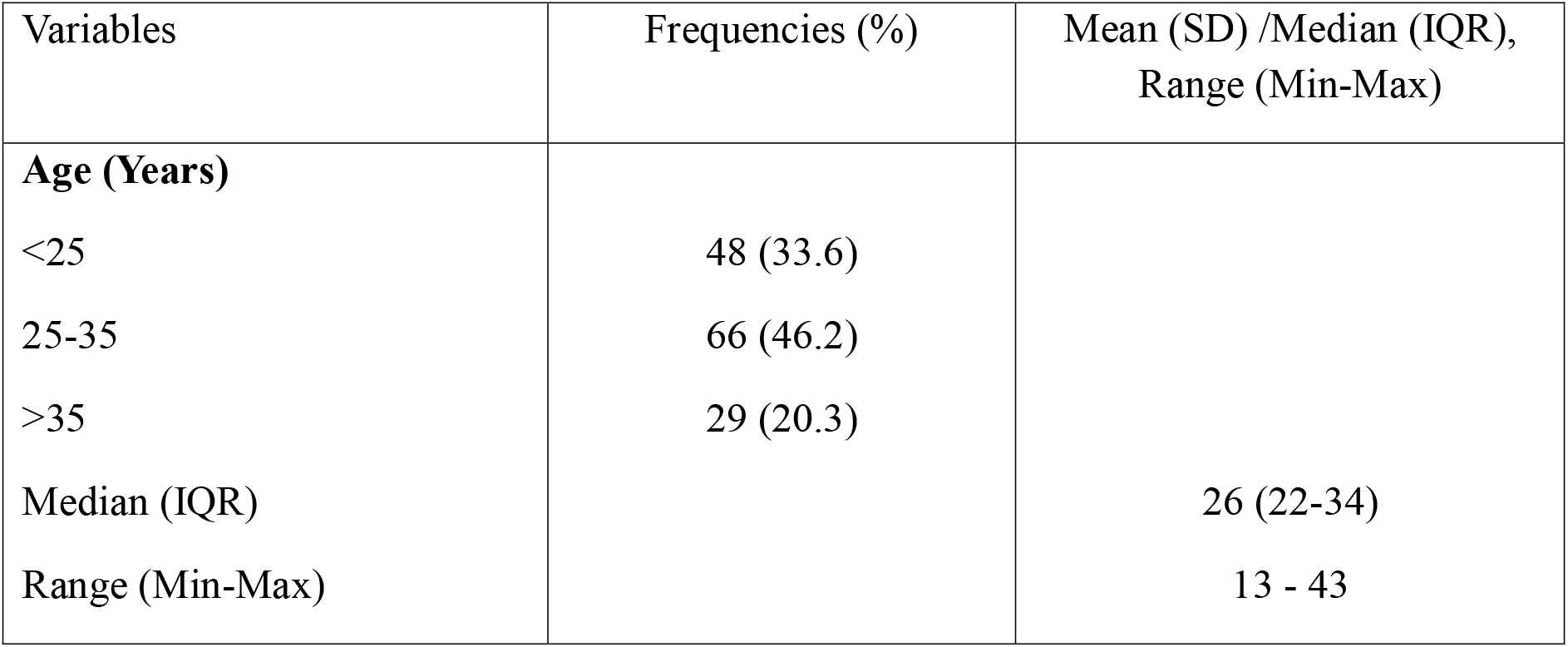
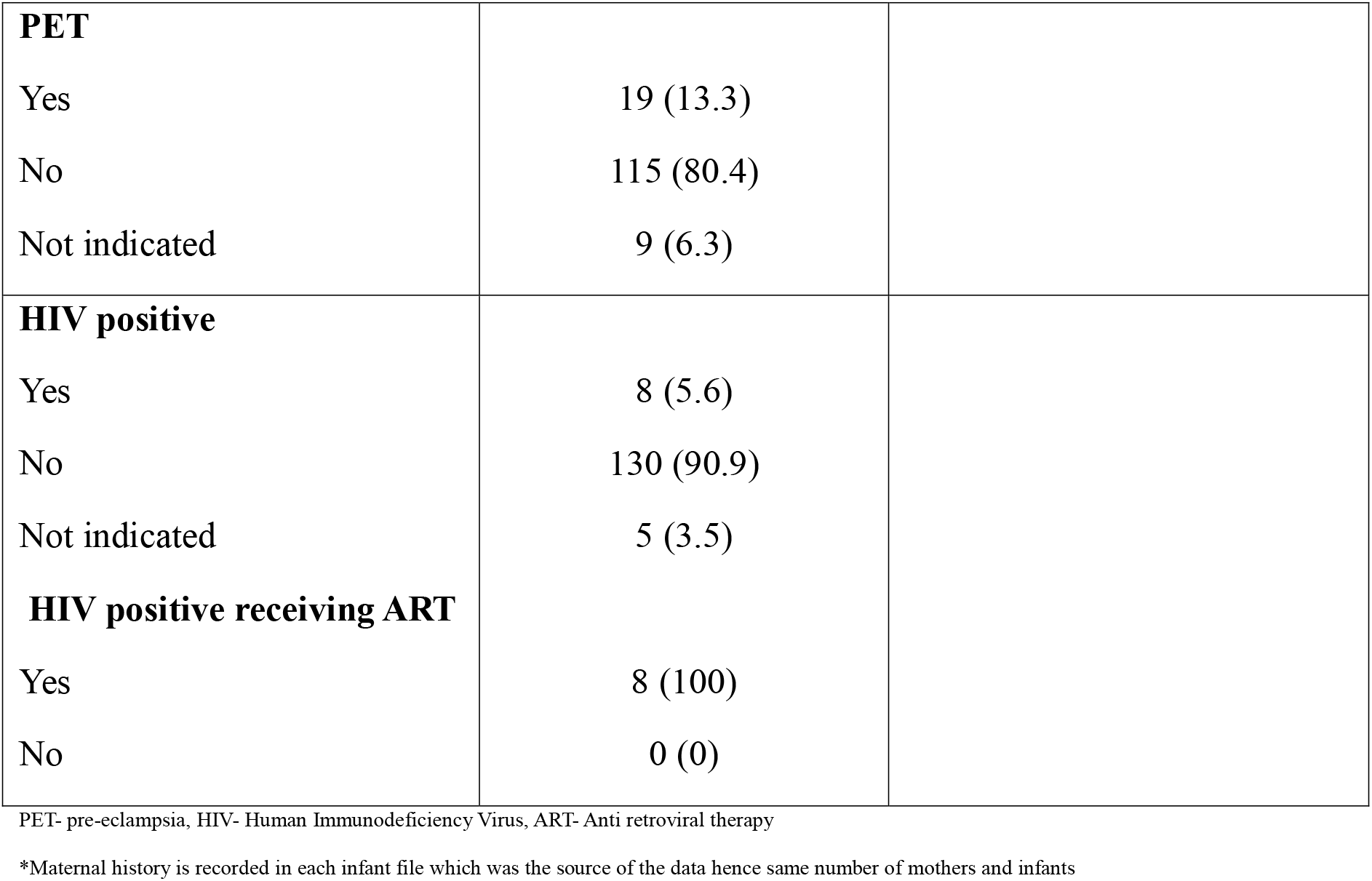
Maternal demographic characteristics (N=143*)

### The nutritional information of the LBW infants studied

The nutritional data extracted included date and time of enteral feed initiation, whether it was breast milk or formula, date of first attaining full enteral feeds, whether the infant received any supplements (multivitamin, iron, folate) and use of human milk fortification. Most of the infants, 86.7%, received their first enteral feed within the first 48 hours after birth with the majority (60.1%) receiving expressed breastmilk. It took seven days or more to achieve full enteral feeds at 180mls/kg/day in majority of the babies (65%) with a median of 8 days (IQR 6-12). None of the infants studied had received any human milk fortification while other supplements (iron, multivitamin, and folate) were not routinely used. The findings are summarized in Table 3 below.

**Table 3:** Nutritional information of the LBW infants (n=143)

| Variable | Frequencies (%) | Mean (SD) /Median (IQR), Range (Min-Max) |
| --- | --- | --- |
| **Time to 1<sup>st</sup> feed (hours)** |  |  |
| <48 | 124 (86.7) |  |
| ≥48 | 19 (13.3) |  |
| Median (IQR) |  | 20.9 (10.9-34.8) |
| Range (Min-Max) |  | 0.37-68 |
| **Time to full feeds**<br>**(180mls/kg/day) (days)** |  |  |
| <7 | 50 (35.0) |  |
| ≥7 | 93 (65.0) |  |
| Median (IQR) |  | 8.0 (6.0-12.0) |
| Range (Min-max) |  | 3–33 |
| **Type of 1<sup>st</sup> feed** |  |  |
| Breastmilk | 86 (60.1) |  |
| Formula | 57 (39.9) |  |
| **Use of supplements** |  |  |
| Iron | 39 (27.3) |  |
| Folate | 57 (39.9) |  |
| Multivitamins | 61 (42.7) |  |

### Clinical characteristics and outcome data of the study subjects

The infants were assessed for some common morbidities during their hospitalization period. The file diagnosis of sepsis and respiratory distress syndrome (RDS) was found to have been made in most of them at 83.9% and 53.9% respectively. The majority, (86%), of the infants had not used continuous positive airway pressure (CPAP), while 68.5% had required supplemental oxygen during their hospitalization. Slightly above three quarters, (78.3%), did not receive blood transfusion. Most, (92.3%), of the infants were discharged alive. The mean time to regain birth weight was 14.2 days (SD: 6.5) with a range of 1-37 days. The median duration of hospital stay was 30 days (IQR: 20-41) with a range of 7-101 days.

These findings are presented in Table 4 below.

**Table 4:** clinical characteristics and outcome data of the LBW infants studied (n=143)

| Variable | Frequency (%) | Mean (SD) /Median (IQR),<br>Range (Min-Max) |
| --- | --- | --- |
| **File diagnosis of sepsis** |  |  |
| Yes | 120 (83.9) |  |
| No | 23 (16.1) |  |
| **File diagnosis of RDS** |  |  |
| Yes | 77 (53.9) |  |
| No | 66 (46.1) |  |
| **Use of CPAP** |  |  |
| Yes | 20 (14) |  |
| No | 123 (86) |  |
| **Any use of supplemental oxygen** |  |  |
| Yes |  |  |
| No | 98 (68.5) |  |
|  | 45 (31.5) |  |
| **Any blood transfusion** |  |  |
| Yes | 31 (21.7) |  |
| No | 112 (78.3) |  |
| **Time to regain birth weight** |  |  |
| 14 days or less | 73 (51.1) |  |
| >14 days | 70 (49.0) |  |
| Mean (SD) |  | 14.2 (6.5) |
| Range (Min-Max) |  | 1-37 |
| **Duration of hospital stay (days)** |  |  |
| ≤ 30 | 72 (50.4) |  |
| >30 | 71 (49.7) |  |
| Median (IQR) |  | 30.0 (20.0-41.0) |
| Range (Min-Max) |  | 7-101 |
| **Outcome** |  |  |
| Discharged | 132 (92.3) |  |
| Transferred to another facility | 2 (1.4) |  |
| Died | 9 (6.3) |  |
| **Exit weight centile (cross sectional PNGR)** |  |  |
| <3<sup>rd</sup> | 64 (44.8) |  |
| 3<sup>rd</sup>-10<sup>th</sup> | 26 (18.2) |  |
| ≥10<sup>th</sup> | 53 (37.1) |  |
| **Exit weight Z scores** |  |  |
| <-2 SDs | 60 (42.0) |  |
| <-1 to -2 SDs | 41 (28.7) |  |
| ≥-1 SDs | 42 (29.4) |  |

### Prevalence of postnatal growth restriction (PNGR) among LBW infants

Postnatal growth restriction (PNGR) was determined as either cross sectional, discharge weight <10^th^ centile, or longitudinal if discharge weight Z score had dropped by >1SD from birth weight. These were further classified as severe cross sectional PNGR if discharge weight was <3^rd^ centile or if change in z score was more than 2SD from birth weight. The LBW infant was flagged as having moderate PNGR if discharge weight was determined to be between 3^rd^-10^th^ centiles or if Z score dropped by 1-2SD for cross sectional and longitudinal PNGR respectively.

The prevalence of PNGR was found to be 63% and 53.2% for the cross sectional and longitudinal classifications respectively. According to cross-sectional and longitudinal criteria, 44.8% and 16.1% of study participants were found to have severe PNGR, while 18.2% and 30.7% showed moderate PNGR, respectively. It was notable that the proportion of infants with weight <10^th^ centile at discharge increased to 63% from 28.7% at birth while those with discharge Z score <-2 increased to 42% from 18.2%. These findings are summarized in Table 5 below.

**Table 5:**
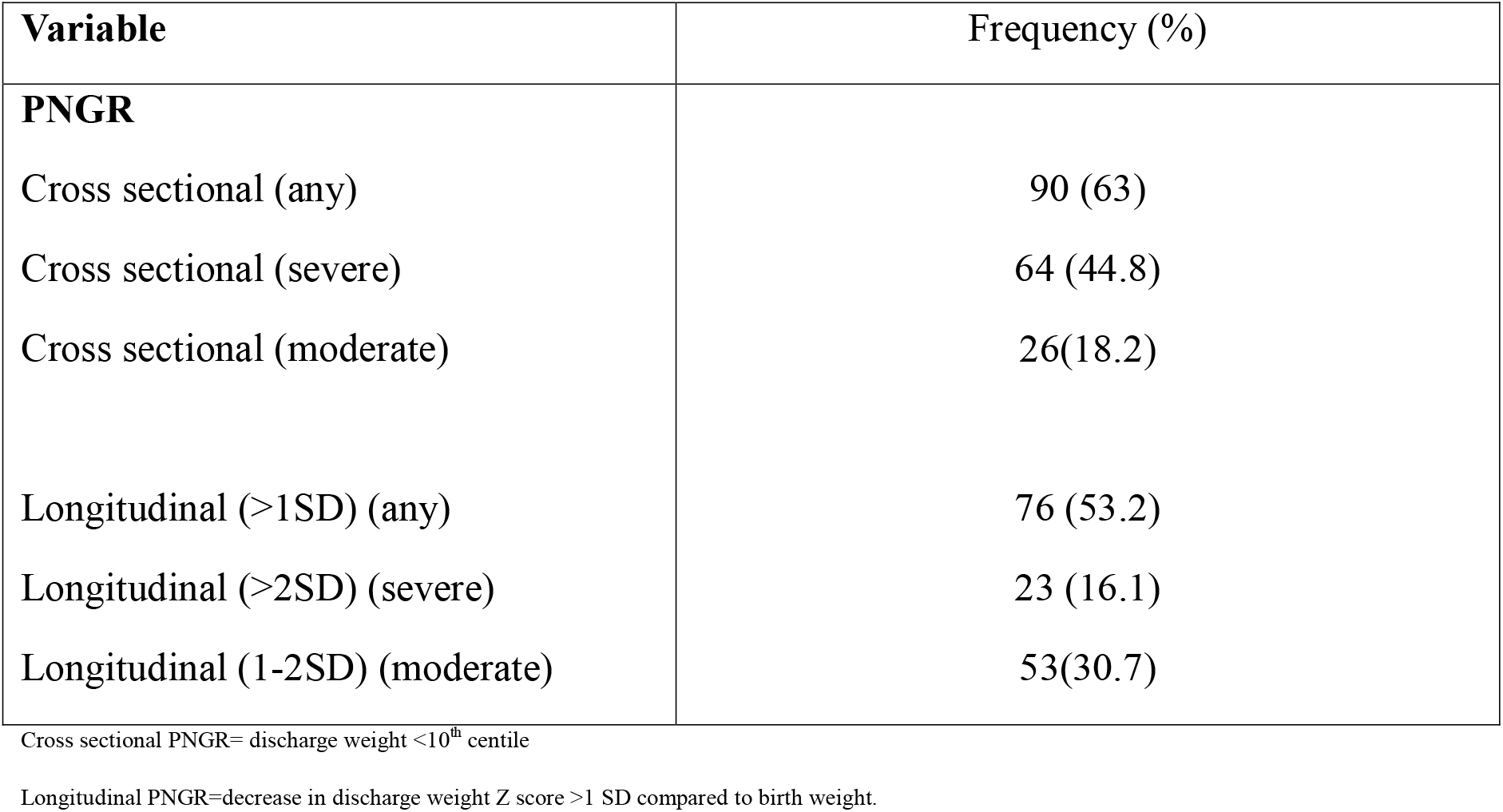
Prevalence of PNGR.

| Variable | Frequency (%) |
| --- | --- |
| **PNGR** |  |
| Cross sectional (any) | 90 (63) |
| Cross sectional (severe) | 64 (44.8) |
| Cross sectional (moderate) | 26(18.2) |
| Longitudinal (>1SD) (any) | 76 (53.2) |
| Longitudinal (>2SD) (severe) | 23 (16.1) |
| Longitudinal (1-2SD) (moderate) | 53(30.7) |

Cross sectional PNGR= discharge weight <10<sup>th</sup> centile
Longitudinal PNGR=decrease in discharge weight Z score >1 SD compared to birth weight.

### Factors associated with PNGR among LBW infants

The infant and maternal characteristics were analyzed to determine their association with cross sectional and longitudinal PNGR among LBW infants <1800g.

### cross-sectional post-natal growth restriction

Maternal age >35 years, female gender, gestation age at birth 32-35 weeks, birth weight <1000g and a 5-minute Apgar score <5 all increased the risk of cross sectional PNGR. The nutritional and clinical characteristics that were found to increase the risk of cross sectional PNGR included 1^st^ feed ≥ 48 hours, breast milk as the 1^st^ enteral feed, ≥ 7 days to achieve full enteral feeds, use of supplements, history of a blood transfusion and hospital stay longer than 30 days. Only gender and gestation age at birth were found to be statistically significant. This analysis is summarized and presented in more detail in Tables 6 and 7 below.

**Table 6:** Bivariate analysis of maternal and infant characteristics and association with Cross-sectional PNGR.

| Variable | Cross sectional PNGR |  |  |  | OR (95% CI) | p-value |
| --- | --- | --- | --- | --- | --- | --- |
|  | Yes |  | No |  |  |  |
| Maternal age (years) | (n=90) | % | (n=53) | % |  |  |
| <25 | 27 | 30.0 | 21 | 39.6 | Reference |  |
| 25 – 35 | 42 | 46.7 | 24 | 45.3 | 1.4 (0.6 – 2.9) | 0.426 |
| >35 | 21 | 23.3 | 8 | 15.1 | 2.0 (0.8 – 5.5) | 0.159 |
| Infant gender |  |  |  |  |  |  |
| Male | 41 | 45.6 | 36 | 67.9 | Reference |  |
| Female | 49 | 54.4 | 17 | 32.1 | 2.5 (1.2 – 5.2) | 0.010 |
| Gestation at birth (weeks) |  |  |  |  |  |  |
| <28 | 4 | 4.4 | 10 | 18.9 | Reference |  |
| 28 – 31 | 23 | 25.6 | 32 | 60.4 | 1.8 (0.5 – 6.4) | 0.369 |
| 32 – 35 | 46 | 51.1 | 11 | 20.8 | 10.5 (2.8 – 39.7) | 0.001 |
| ≥36 | 17 | 18.9 | 0 | 0.0 | - | - |
| **Birth weight grams** |  |  |  |  |  |  |
| <1000 | 9 | 10.0 | 3 | 5.7 | Reference |  |
| 1000 – 1499 | 36 | 40.0 | 30 | 56.6 | 0.4 (0.1 – 1.6) | 0.198 |
| ≥1500 | 45 | 50.0 | 20 | 37.7 | 0.8 (0.2 – 3.1) | 0.689 |
| **Birth weight centile** |  |  |  |  |  |  |
| <3<sup>rd</sup> | 29 | 32.2 | 0 | 0.0 | - | - |
| 3<sup>rd</sup> -<10<sup>th</sup> | 12 | 13.3 | 0 | 0.0 | - | - |
| ≥10<sup>th</sup> | 49 | 54.4 | 53 | 0.0 | Reference |  |
| **Apgar score at 5 minutes** |  |  |  |  |  |  |
| <5 | 11 | 12.4 | 4 | 7.5 | 1.7 (0.5 – 5.7) | 0.367 |
| ≥5 | 78 | 87.6 | 49 | 92.5 | Reference |  |

OR – Odds Ratio, CI – Confidence Interval, PNGR-post natal growth restriction

**Table 7:**
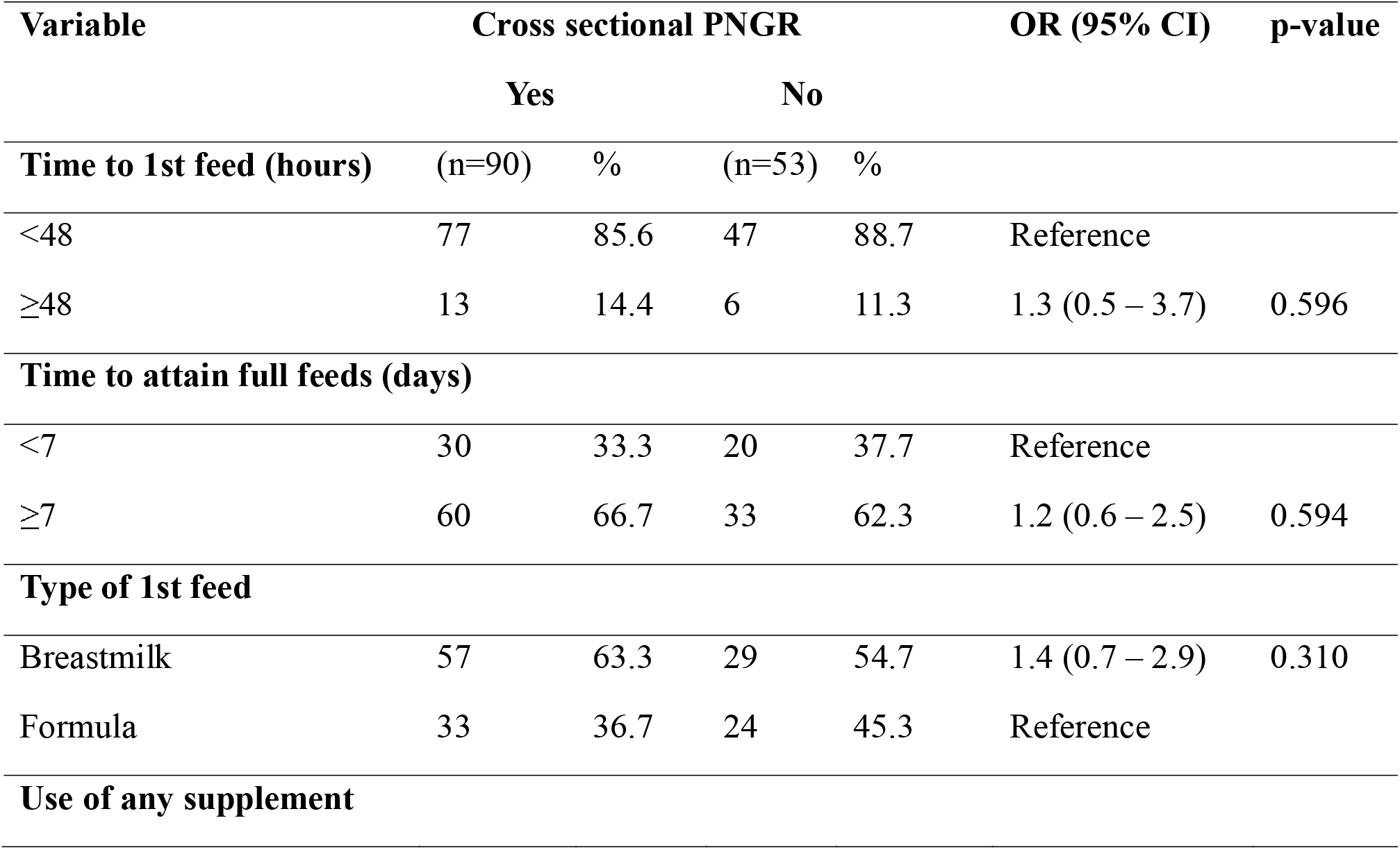
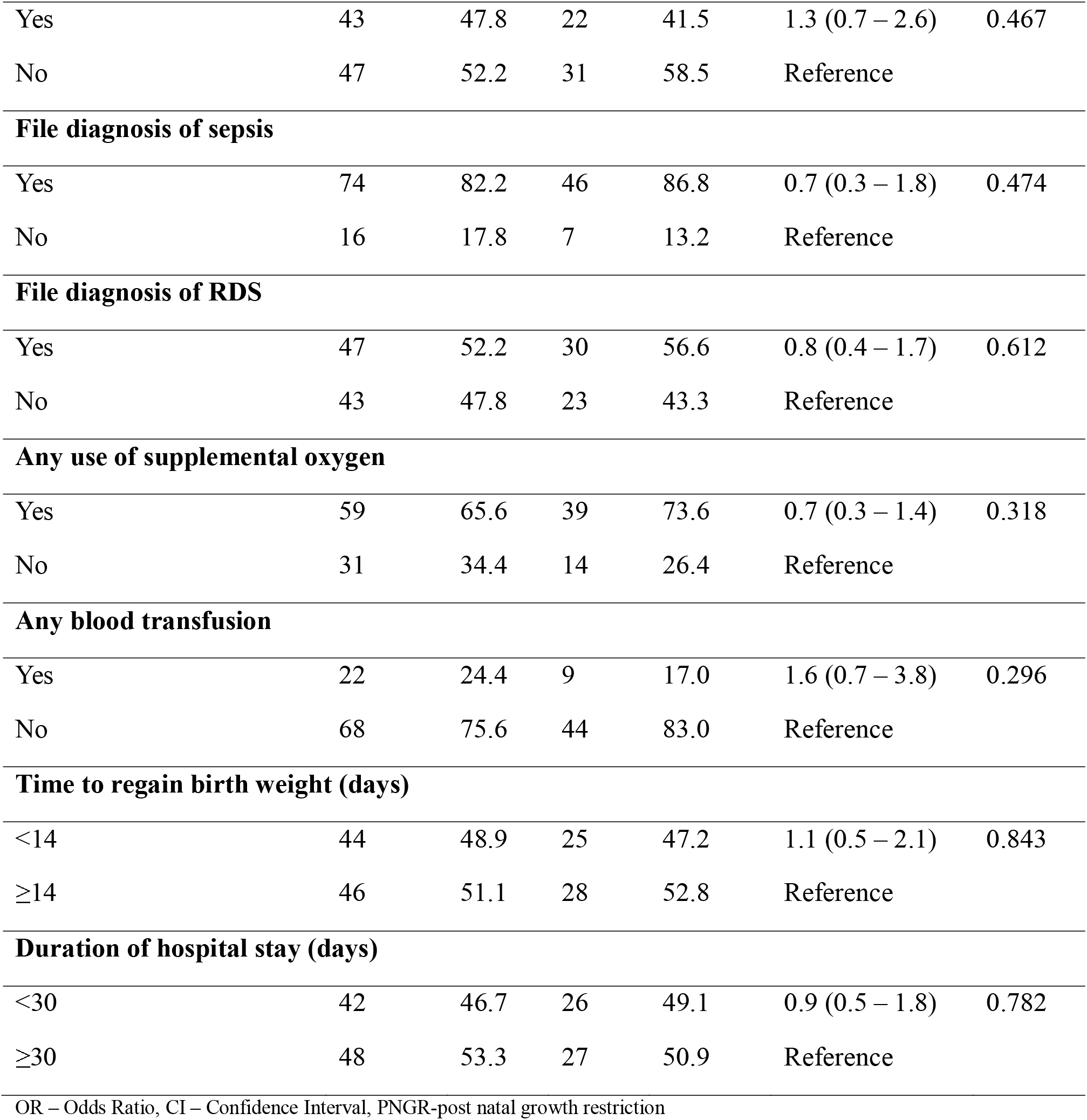
Bivariate analysis of nutritional, clinical and outcome characteristics and association with Cross-sectional PNGR.

### Longitudinal post-natal growth restriction

Longitudinal PNGR was found to be increased, in younger maternal age <25 years, male gender, gestation at birth <28 weeks, birth weight <1000g, birth weight <3^rd^ centile and 5-minute Apgar score <5. In the analysis of clinical and nutritional characteristics, 1^st^ feed >48 hours, breast milk as the 1^st^ enteral feed, achieving full feeds >7days, having received supplements, a diagnosis of sepsis, RDS, use of supplemental oxygen, having received a blood transfusion, > 14 days to regain birth weight and >30 days of hospital stay all increased the risk of longitudinal PNGR. This is presented in tables 8 and 9 below.

**Table 8:**
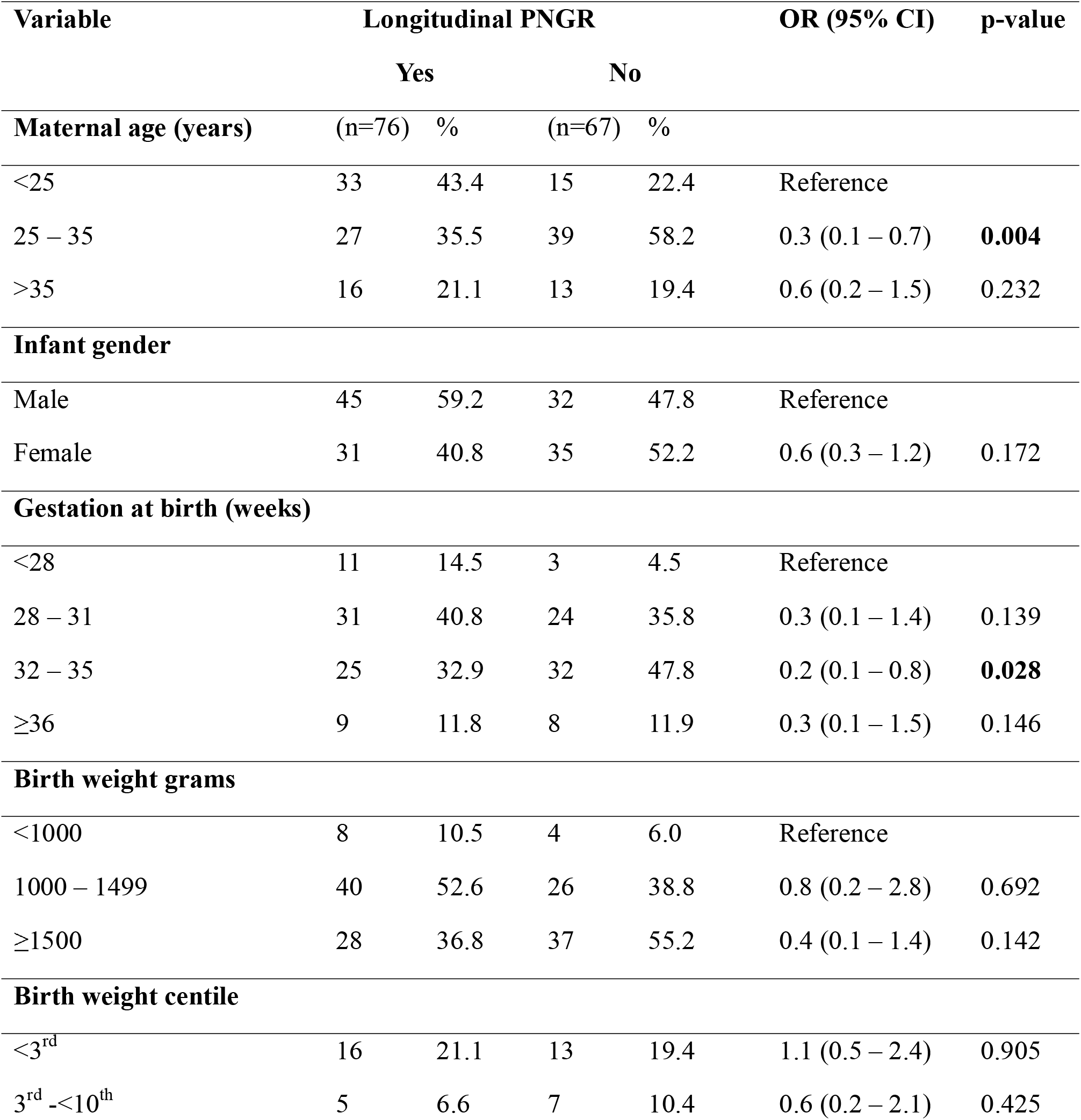
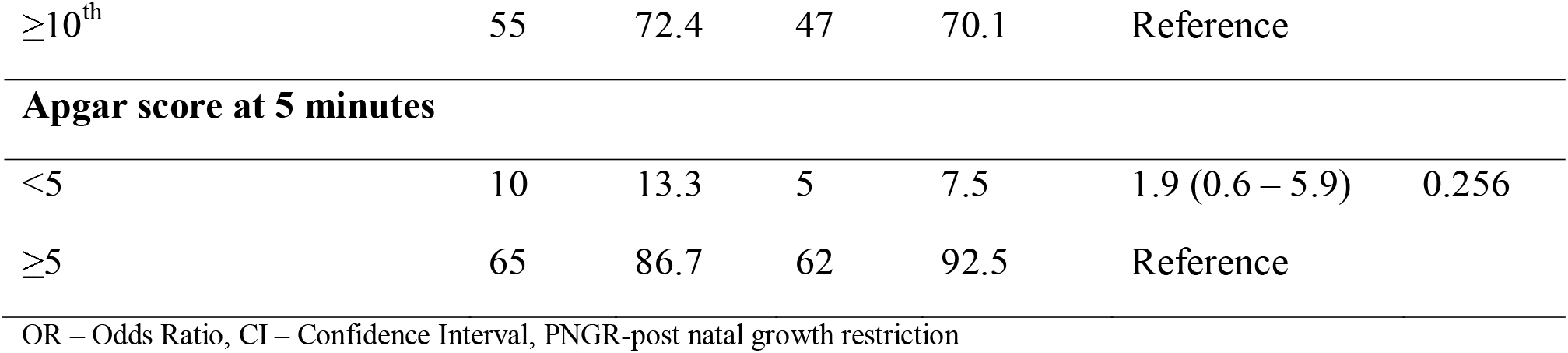
Bivariate analysis of maternal and infant characteristics and association with Longitudinal PNGR.

| Variable | Longitudinal PNGR |  |  |  | OR (95% CI) | p-value |
| --- | --- | --- | --- | --- | --- | --- |
|  | Yes |  | No |  |  |  |
| Maternal age (years) | (n=76) | % | (n=67) | % |  |  |
| <25 | 33 | 43.4 | 15 | 22.4 | Reference |  |
| 25 – 35 | 27 | 35.5 | 39 | 58.2 | 0.3 (0.1 – 0.7) | **0.004** |
| >35 | 16 | 21.1 | 13 | 19.4 | 0.6 (0.2 – 1.5) | 0.232 |
| Infant gender |  |  |  |  |  |  |
| Male | 45 | 59.2 | 32 | 47.8 | Reference |  |
| Female | 31 | 40.8 | 35 | 52.2 | 0.6 (0.3 – 1.2) | 0.172 |
| Gestation at birth (weeks) |  |  |  |  |  |  |
| <28 | 11 | 14.5 | 3 | 4.5 | Reference |  |
| 28 – 31 | 31 | 40.8 | 24 | 35.8 | 0.3 (0.1 – 1.4) | 0.139 |
| 32 – 35 | 25 | 32.9 | 32 | 47.8 | 0.2 (0.1 – 0.8) | **0.028** |
| ≥36 | 9 | 11.8 | 8 | 11.9 | 0.3 (0.1 – 1.5) | 0.146 |
| Birth weight grams |  |  |  |  |  |  |
| <1000 | 8 | 10.5 | 4 | 6.0 | Reference |  |
| 1000 – 1499 | 40 | 52.6 | 26 | 38.8 | 0.8 (0.2 – 2.8) | 0.692 |
| ≥1500 | 28 | 36.8 | 37 | 55.2 | 0.4 (0.1 – 1.4) | 0.142 |
| Birth weight centile |  |  |  |  |  |  |
| <3<sup>rd</sup> | 16 | 21.1 | 13 | 19.4 | 1.1 (0.5 – 2.4) | 0.905 |
| 3<sup>rd</sup> -<10<sup>th</sup> | 5 | 6.6 | 7 | 10.4 | 0.6 (0.2 – 2.1) | 0.425 |
| $\geq 10^{\text{th}}$ | 55 | 72.4 | 47 | 70.1 | Reference | |
| Apgar score at 5 minutes |  |  |  |  |  |  |
| <5 | 10 | 13.3 | 5 | 7.5 | 1.9 (0.6 – 5.9) | 0.256 |
| $\geq 5$ | 65 | 86.7 | 62 | 92.5 | Reference | |

OR – Odds Ratio, CI – Confidence Interval, PNGR-post natal growth restriction

**Table 9:** Bivariate analysis of nutritional, clinical and outcome characteristics and association with Longitudinal PNGR.

| Variable | Longitudinal PNGR |  |  |  | OR (95% CI) | p-value |
| --- | --- | --- | --- | --- | --- | --- |
|  | Yes |  | No |  |  |  |
| Time to 1st feed (hours) | (n=76) | % | (n=67) | % |  |  |
| <48 | 64 | 84.2 | 60 | 89.6 | Reference |  |
| ≥48 | 12 | 15.8 | 7 | 10.4 | 1.6 (0.6 – 4.4) | 0.351 |
| Time to attain full feeds (days) |  |  |  |  |  |  |
| <7 | 20 | 26.3 | 30 | 44.8 | Reference |  |
| ≥7 | 56 | 73.7 | 37 | 55.2 | 2.3 (1.1 – 4.6) | 0.022 |
| Type of 1st feed |  |  |  |  |  |  |
| Breastmilk | 56 | 73.7 | 30 | 44.8 | 3.5 (1.7 – 7.0) | <0.001 |
| Formula | 20 | 26.3 | 37 | 55.2 | Reference |  |
| Use of any supplement* |  |  |  |  |  |  |
| Yes | 49 | 64.5 | 16 | 23.9 | 5.8 (2.8 – 12.0) | <0.001 |
| No | 27 | 35.5 | 51 | 76.1 | Reference |  |
| File diagnosis of sepsis |  |  |  |  |  |  |
| Yes | 68 | 89.5 | 52 | 77.6 | 2.5 (1.0 – 6.2) | 0.054 |
| No | 8 | 10.5 | 15 | 22.4 | Reference |  |
| File diagnosis of RDS |  |  |  |  |  |  |
| Yes | 53 | 69.7 | 24 | 35.8 | 4.1 (2.1 – 8.3) | <0.001 |
| No | 23 | 30.3 | 43 | 64.2 | Reference |  |
| **Any use of supplemental oxygen** |  |  |  |  |  |  |
| Yes | 59 | 77.6 | 39 | 58.2 | 2.5 (1.2 – 5.1) | **0.013** |
| No | 17 | 22.4 | 28 | 41.8 | Reference |  |
| **Any blood transfusion** |  |  |  |  |  |  |
| Yes | 26 | 34.2 | 5 | 7.5 | 6.4 (2.3 – 18.0) | **<0.001** |
| No | 50 | 65.8 | 62 | 92.5 | Reference |  |
| **Time to regain birth weight (days)** |  |  |  |  |  |  |
| <14 | 17 | 22.4 | 52 | 77.6 | 0.1 (0.03 – 0.2) | **<0.001** |
| ≥14 | 59 | 77.6 | 15 | 22.4 | Reference |  |
| **Duration of hospital stay (days)** |  |  |  |  |  |  |
| <30 | 20 | 26.3 | 48 | 71.6 | 0.1 (0.1 – 0.3) | **<0.001** |
| ≥30 | 56 | 73.7 | 19 | 28.4 | Reference |  |

OR – Odds Ratio, CI – Confidence Interval, PNGR-post natal growth restriction

The infant, maternal and clinical characteristics with a p-value of <0.05 in the bivariate analysis of the longitudinal PNGR were subjected to a multivariate logistic regression model.

The use of any nutritional supplement, iron, folate or multi-vitamins, was interestingly found to increase the odds of longitudinal PNGR aOR 3.0 (95% CI 1.1-8.3). The infants who regained their birth weight in less than 14 days aOR 0.1 (95% CI 0.1-0.4) and had been admitted for less than 30 days aOR 0.3 (95% CI 0.1-0.9) had less risk of longitudinal PNGR. These three factors remained statistically significant at multivariate analysis. Table 10 below presents a summary of this analysis.

**Table 10:** Multivariate analysis of nutritional, clinical and outcome characteristics and association with Longitudinal PNGR.

| Variable | cOR (95% CI) | p-value | aOR (95% CI) | p-value |
| --- | --- | --- | --- | --- |
| **Time to attain full feeds (days)** |  |  |  |  |
| <7 | Reference |  | Reference |  |
| ≥7 | 2.3 (1.1 – 4.6) | **0.022** | 1.6 (0.6 – 4.2) | 0.327 |
| **Type of 1st feed** |  |  |  |  |
| Breastmilk | 3.5 (1.7 – 7.0) | **<0.001** | 1.3 (0.5 – 3.5) | 0.601 |
| Formula | Reference |  | Reference |  |
| **Use of any supplement** |  |  |  |  |
| Yes | 5.8 (2.8 – 12.0) | **<0.001** | 3.0 (1.1 – 8.3) | **0.037** |
| No | Reference |  | Reference |  |
| **File diagnosis of RDS** |  |  |  |  |
| Yes | 4.1 (2.1 – 8.3) | **<0.001** | 1.5 (0.5 – 5.1) | 0.503 |
| No | Reference |  | Reference |  |
| **Any use of supplemental oxygen** |  |  |  |  |
| Yes | 2.5 (1.2 – 5.1) | **0.013** | 0.6 (0.2 – 2.1) | 0.407 |
| No | Reference |  | Reference |  |
| **Any blood transfusion** |  |  |  |  |
| Yes | 6.4 (2.3 – 18.0) | **<0.001** | 1.7 (0.5 – 6.1) | 0.423 |
| No | Reference |  | Reference |  |
| **Time to regain birth weight (days)** |  |  |  |  |
| <14 | 0.1 (0.03 – 0.2) | **<0.001** | 0.1 (0.1 – 0.4) | **<0.001** |
| ≥14 | Reference |  | Reference |  |
| **Duration of hospital stay (days)** |  |  |  |  |
| <30 | 0.1 (0.1 – 0.3) | **<0.001** | 0.3 (0.1 – 0.9) | **0.025** |
| ≥30 | Reference |  | Reference |  |

cOR- crude odds ratio
aOR- adjusted odds ratio

substantial multicollinearity was observed between diagnosis of RDS and use of any supplemental oxygen, as well as time to regain birth weight and the duration of hospital stay. When adjusted for each other, due to their close clinical relation, the statistical significance of all the other variables except duration of hospital stay and time to regain birth weight was lost.

## Discussion

Significant measures have been put in place to enhance the survival rates of low-birth-weight neonates. It is thus important to ensure that this surviving neonate thrives as well. Post-natal growth restriction (PNGR) has been associated with long term morbidity including poor neurodevelopmental and metabolic outcomes that persist in adulthood. It is therefore important to determine the burden of PNGR among our surviving LBW infants using locally generated data. Postnatal growth restriction (PNGR) has been suggested in other studies to be universal in preterm and LBW infants (3). There are two ways to define PNGR, longitudinal (drop in Z score by >1SD from birth to discharge weight) and cross sectional (discharge weight below the 10th centile) (19, 15).

This was a retrospective single hospital study, which as far as we know, is the first of its kind in Kenya to assess postnatal growth after initial hospitalization in neonates. We used a cut of weight of <1800g since this is the approximate target weight for discharge from the Nyeri County referral hospital (NCRH) newborn unit (NBU). Most published studies use birth weight <1500g for recruitment (3, 4 10,13). We restricted ourselves from using a gestation age for study recruitment since most mothers in our set up are usually unsure of their last menstrual period date and there are very few, if any, who get early obstetric ultrasounds that are more accurate in dating a pregnancy. The study site also does not routinely do Ballard scoring to confirm gestation of the preterm and LBW infants admitted. This is different from most available studies that use <32 weeks and a few that use 28 weeks gestation as a cut off for study recruitment. While our study findings were comparable to most similar studies, our participant selection that included bigger and likely more mature babies makes the need to do the comparison cautiously.

### Cross-sectional post-natal growth restriction

We found that among all the infants we studied, 90 (63%) had cross sectional PNGR. It was notable in our study that the proportion of infants with weight <10^th^ centile at discharge increased to 63% from 28.7% at birth while the discharge Z score <-2 SD increased to 42% from 18.2% at birth. This observation is like what was observed in a Brazilian 4-unit study which observed that 11% of their study subjects had IUGR at birth and this rate of growth restriction increased to 77.4% at discharge considering weight (15). This prevalence was significantly higher than 21.7% found in a Boston study (6), 51% and 43% extra uterine growth restriction rate of the SGA infants and non-SGA infants respectively < 32 weeks studied in Barcelona, Spain (14) and 42% finding of the Vermont Oxford Network (16). The prevalence (63%) was however much less than 84% found in a Hong Kong neonatal unit 10-year study of neonatal outcomes (16), 91.2% findings in a South African single center study (17), 89% in a Nigerian study (18) and 73% in a rural Ugandan Study (19).

Our study found 44.8% (<3^rd^ centile discharge weight) severe cross sectional PNGR which was considerably lower than the 79.6% finding in a South African study (17), and 77% in a Nigerian multicenter study (19).

Previous studies have found that infants who are born SGA have the greatest incidence of PNGR. Among our study subjects, 28.6% were SGA at birth. All these infants had cross-sectional PNGR at discharge, consistent with a West Indian study where infants born at <3rd centile from 34-37 weeks gestation maintained that growth to term corrected gestation (13) and a Spanish study that found EUGR among 51% of SGA infants compared to 43% of non-SGA infants at discharge (14). Similar effects of SGA were observed in a Brazilian study that showed 54.2% (p=0.000) of SGA infants had EUGR at discharge compared to 12.3% AGA infants (15).

### Longitudinal post-natal growth restriction

Longitudinal PNGR was observed in 76 (53.2%) of our study subjects which was much higher than 36.2% found in a 2-center study in Southeast Asia (10), 26% found in 4 neonatal units’ study done in Rio, Brazil (15), an Israeli study that found overall no PNGF among 74.2% of VLBW infants with no major comorbidity vs 45.5% of those with one major comorbidity (4) and lower than a West Indian study that found all VLBW infants studied dropped centile lines for weight from birth to 40 weeks PMA (13).

In terms of severity, 16.1% (drop by >2SD from birth to discharge Z score for weight) severe PNGR was observed in our study subjects. This finding was lower than the overall 28.8% and 36.5% severe post-natal growth faltering (change in Z score >2SD at 36 weeks corrected gestation) for VLBW and ELBW infants respectively in a Southeast Asian study (10). Our findings were comparable to 13.6% severe PNGF among infants with one major comorbidity in an Israeli study but significantly higher than 2.1% observed in infants with no major comorbidity in the same study (4).

Approximately half of the infants with SGA at birth in our study were found to have longitudinal PNGR at discharge. In Brazil, they found that being SGA at birth increased by 4.33 times the chance of being growth restricted at discharge (15) same as in Barcelona where EUGR was observed in 51% vs 43% among SGA vs non-SGA respectively (14). This effect of SGA at birth on PNGR was replicated in other studies, (12, 20).

These findings underscore the significant prevalence of PNGR, both longitudinal and cross-sectional, in NCRH NBU graduates. These differences in prevalence could be explained by the regional differences in newborn care practices, available infrastructure and feeding protocols.

### Factors associated with PNGR

Clinical conditions such as sepsis (OR 2.5 (1-6.2)), respiratory distress syndrome (RDS) (OR 4.1(2.1-8.3), and the need for any supplemental oxygen (OR 2.5 (1.2-5.1)) prevalent among our studied infants demonstrated strong associations with longitudinal PNGR. We found that a diagnosis of sepsis (OR=0.7), was oddly associated with reduced risk of cross-sectional PNGR but showed increased odds for longitudinal PNGR OR 2.5 (1.2-5.1). Sepsis was found to be positively associated with PNGR in a West Indian study that found that 65% of culture positive sepsis patients had PNGR at discharge (13) similar to a Ugandan study that found a significant association AOR 6.76 (95% CI 2.15-21.2) between sepsis diagnosis and PNGR (19). A South African study however found that despite 45.5% of study subjects having blood culture confirmed sepsis, there was no difference in growth compared to those without sepsis (13,17).

Other neonatal morbidities that have been strongly associated with PNGR include BPD, severe IVH, PDA and NEC (4,6,12,19). The impact of neonatal morbidity on post-natal growth was similarly demonstrated in an Israeli study that showed 57.5% and 25.8% had PNGR among those with and those without any major morbidity respectively (4). In our study, RDS (OR=0.8) and use of supplemental oxygen (OR=0.7) were surprisingly associated with reduced odds of cross sectional PNGR while the reverse effect was observed for longitudinal PNGR at OR=4.1 and 2.5 respectively. A study in Rio, Brazil found that a diagnosis of RDS reduced by 35% the chance of growth restriction, based on weight, by discharge (15). The same study, however, noted that the RDS diagnosis was more common among appropriate for gestation (AGA) infants. In an Israeli study, RDS had a greater effect on PNGF in infants without other morbidities, with ORs of 3.4 for severe and 1.84 for mild PNGF vs no PNGF (p<0.001) (4). The infants who received a blood transfusion however showed increased risk of both longitudinal (OR= 6.4) and cross-sectional (OR=1.6) PNGR. Our study was, however, unable to demonstrate PNGR association with these specific morbidities (PDA, BPD, IVH, NEC) since the number of patients with such diagnosis was too few to be analyzed.

We found that a 5-minute Apgar score of <5 was associated with an increased risk (OR 1.7 (0.5-5.7) of cross-sectional and longitudinal OR 1.9 (0.6-5.9) PNGR respectively. This finding was comparable to the British birth cohort study that found on average lower Apgar scores for babies who were discharged SGA (6).

The importance of early initiation of enteral feeds even during parenteral nutrition has been demonstrated in previous studies. We found an increased risk of longitudinal PNGR (OR 1.6 (0.6-4.4)) and cross sectional PNGR (OR 1.3 (0.5-3.7) among infants who received their first enteral feed beyond 48 hours after birth. A British birth cohort study found that delayed initiation of enteral feeds increased the odds of PNGR (OR 1.04, p = 0.001, CI 1.01–1.06) (6). A Ugandan study concluded that delayed initiation of enteral feeds beyond 48 hours was responsible for PNGR (19). We also observed an increased risk of PNGR in infants who received breast milk as their first enteral feed (OR 1.4 (0.7-2.9)) for cross sectional and OR 3.5(1.7-7.0) p<0.01 for longitudinal PNGR. This was mirrored in a Nigerian study that found more PNGR in infants who received breast milk (20). Most units appreciate the benefits of mother’s own milk, which is highly recommended for all neonates including preterms and LBW infants, and may choose to wait for this to be available before starting enteral feeds. unavailability of parenteral nutrition to support the high nutritional needs early on, coupled with the unavailability of mother’s own milk or donor human milk, lack of formula or unwillingness to give formula in a sick neonate may likely have contributed to PNGR seen in infants who received breast milk as the first feed rather than a direct effect of the breast milk feed.

The delayed attainment of full enteral feeds which was found in 65% of our study participants has been linked to PNGR (6). We observed that a duration longer than 7 days to achieve full enteral feeds increased the odds ratio of both longitudinal (OR 2.3 (1.1-4.6)) and cross sectional (OR 1.2(0.6-2.5) PNGR. Previous studies have shown that rapid advancement to full enteral feeds at about 1 week is associated with better growth in preterm and LBW infants (11, 17,19). The importance of early parenteral nutrition and breast milk fortification including use of supplements has been underscored in several previous studies (10, 11, 23, 24, 25). Despite our high prevalence of PNGR, we observed that most (86.7%) of our subjects received their first enteral feed before 48 hours as recommended. This could point to the importance of meeting the high protein and caloric requirements for LBW infants’ growth that cannot be met by breast milk alone. Our study site does not practice any parenteral nutrition, beyond 10% dextrose solution, no human milk fortification, and the use of supplements is inconsistent. Only 45% of our study population had received any of the three recommended supplements (iron, folate or multivitamins) during their initial hospitalization. This can be attributed to lack of infrastructure, to provide total parenteral nutrition, and poor socioeconomic status that prevents affordability of both supplements and human milk fortifiers. It is interesting to note that we found an increased risk of longitudinal PNGR (OR 5.8(2.8-12) which was statistically significant, and this significance was maintained at multivariate analysis with (aOR 3 (1.1-8.3) (p 0.037)) in infants who received any supplement. This could be attributed to the supplements being given to babies already demonstrating poor growth rather than the recommended routine supplementation.

We observed that hospital stays of 30 days or more significantly increased the risk of longitudinal PNGR (p<0.001), a finding confirmed at multivariate analysis. This finding mirrors other studies where duration of hospital stays, which has been suggested to be a marker of the morbidity burden on the neonate, was associated with PNGR (3,15,19,20). A Brazilian study found that adding one day to hospitalization time increased by 2% the chance of growth restriction at discharge (15). Babies who took 14 days or more to regain their birth weight showed increased risk of having longitudinal PNGR OR 0.1(0.03-0.2 (p<0.001)) and this statistical significance was maintained at multivariate analysis (aOR 0.1(0.1-0.3) (p<0.001)). This is likely because sicker babies face more feeding challenges, grow slower, and will have a longer hospital stay especially in a center like ours without parenteral nutrition or human milk fortification to meet their high nutritional needs.

Young maternal age <25 years was found to be associated with longitudinal PNGR in our study. This could be postulated to be due to low socio-economic status impacting access to antenatal care or poor maternal nutritional status which can be explored by further studies. Maternal pregnancy related complications like pre-eclampsia, anemia and diabetes have been linked to PNGR but we could not analyze the same due to the few numbers with the morbidities in our study. The use of antenatal corticosteroids could also not be studied since it was not routinely recorded in the files.

We observed that longitudinal PNGR, which monitors changes in an infant’s growth centile over time, demonstrated more significant associations with clinical factors than cross-sectional PNGR, which is based solely on a single measurement at discharge. Longitudinal PNGR is often considered a more accurate reflection of growth impairment because it captures dynamic changes and trends in growth patterns, identifying infants who experience a drop in growth centiles during their hospital stay—even if their final measurement remains above the 10th percentile. For example, infants who are born SGA and remain below 10^th^ centile, as well as those who start above the 10th centile but decline over time without reaching <10^th^ centile, may not be identified by cross-sectional methods alone. Therefore, using longitudinal PNGR enables clinicians to recognize a broader group of at-risk infants for closer monitoring and intervention, rather than overlooking those whose risk would be missed by cross-sectional criteria alone.

## Conclusion and recommendations

In conclusion, more than half of all the LBW infants discharged from NCRH NBU have PNGR whether longitudinal or cross sectional. Duration of 14 days or more to regain birth weight, initial admission of 30 days or more and having received any supplement (iron, folate or multivitamin) were significantly associated with longitudinal PNGR.

This study establishes baseline local data that may inform the design of a broader multicenter investigation on PNGR. Such efforts can help pinpoint opportunities for enhancement and support the development of subsequent quality improvement initiatives in neonatal nutrition. Some studies done in low resource settings have concluded that supporting the growth of preterm infants requires feed fortification and micronutrient supplementation especially in these settings that have high incidents of malnutrition and stunting later in life (29). Efforts must be put in place, including but not limited to, resource allocation to provide nutrition with high protein and calories as recommended for preterm and LBW infants through early parenteral nutrition, breast milk fortification and consistent use of supplements. These efforts may include collaborating with hospital administration to secure funding for human milk fortifiers and supplements, partnering with county and national governments to establish regional breast milk banks and encouraging local manufacturers to produce affordable parenteral nutrition options can help ensure that essential nutrients are accessible even in resource-limited settings. By engaging multiple stakeholders and implementing these targeted strategies, we can address the systemic barriers to optimal neonatal nutrition and improve health outcomes for these at-risk infants.

## Study limitations

1. Inherent limitations of the retrospective study design could not be avoided entirely. We were also unable to trace 15 files that could have contributed to the numbers available for the study.
2. This was a single center study whose findings may not necessarily be generalizable. Our selection of study subjects excluded the neonates who died before seven days who may have been sicker introducing a survivorship bias as the sicker neonates are likely underrepresented
3. Our study used a cut-off weight <1800g for enrollment, this means relatively mature and bigger neonates were represented in the study unlike most studies which use gestational age cut-off or mostly <1500g for enrollment. This means the comparisons we made were not entirely direct.Using the patient files for our data extraction assumed that what was documented was accurate and complete

## Data Availability

all data produced in the present work are contained in the manuscript

## Acknowledgements

We appreciate the assistance of the health records information officer at NCRH NBU Ms. Anne Gitonga for her assistance in obtaining the patients’ records. We are grateful to Mr. Wycliffe Ayieko for his assistance during data analysis.

## Author Contributions

**Conceptualization**:Lucy Lyanda

**Data curation**: Lucy Lyanda

**Formal analysis**: Lucy Lyanda, Alfred Keter

**Resources**: Lucy Lyanda

**Methodology**: Lucy Lyanda, Alfred Keter

**Supervision:** Roseline Ochieng’

**Validation:** Roseline Ochieng’

**Visualization:** Lucy Lyanda

**Writing – original draft**: Lucy Lyanda

**Writing – review & editing**: Lucy Lyanda, Roseline Ochieng, Alfred Keter

## Notes

### Competing Interest Statement

The authors have declared no competing interest.

### Author Declarations

The study was approved by the Aga Khan Institutional scientific and ethics review committee (ISERC) ISREC 2024/ISERC-24(v1) who waived informed consent for data access from medical records. A research permit Ref No. 891164 was also obtained from the National Commission for Science, Technology & Innovation (NACOSTI). Additional approvals were sought and granted from the Nyeri County department of health services and the Nyeri county referral hospital administration.

